# Contributions of PTSD polygenic risk and environmental stress to suicidality in preadolescents

**DOI:** 10.1101/2021.05.30.21258082

**Authors:** Nikolaos P Daskalakis, Laura M Schultz, Elina Visoki, Tyler M Moore, Stirling T Argabright, Nathaniel G Harnett, Grace E DiDomenico, Varun Warrier, Laura Almasy, Ran Barzilay

**Author notes:** Corresponding Author: Ran Barzilay, MD, PhD, 10th floor, Gates Pavilion, Hospital of the University of Pennsylvania, 34th and Spruce Street, Philadelphia, PA 19104.

## Abstract

Suicidal ideation and attempts (i.e., suicidality) are complex behaviors driven by environmental stress, genetic susceptibility, and their interaction. Preadolescent suicidality is a major health problem with rising rates, yet its underlying biology is understudied. Here we studied effects of genetic stress susceptibility, estimated by polygenic risk score (PRS) for post-traumatic-stress-disorder (PTSD), on preadolescent suicidality in participants from the Adolescent Brain Cognitive Development (ABCD) Study®. We further evaluated PTSD-PRS effects on suicidality in the presence of environmental stressors that are established suicide risk factors. Analyses included both European and African ancestry participants using PRS calculated based on summary statistics from ancestry-specific genome-wide association studies. In European ancestry participants (N=4,619, n=378 suicidal), PTSD-PRS was associated with preadolescent suicidality (odds ratio [OR]=1.12, 95%CI 1-1.25, p=0.038). Results in African ancestry participants (N=1,334, n=130 suicidal) showed a similar direction but were not statistically significant (OR=1.21, 95%CI 0.93-1.57, p=0.153). Sensitivity analyses using non-psychiatric polygenic score for height and using cross-ancestry PTSD-PRS did not reveal any association with suicidality, supporting the specificity of the association of ancestry-specific PTSD-PRS with suicidality. Environmental stressors were robustly associated with suicidality across ancestries with moderate effect size for negative life events and family conflict (OR 1.27-1.6); and with large effect size (OR ∼ 4) for sexual-orientation discrimination. When combined with environmental factors, PTSD-PRS showed marginal additive effects in explaining variability in suicidality, with no evidence for G X E interaction. Results support use of cross-phenotype PRS, specifically stress-susceptibility, as a robust genetic marker for suicidality risk early in the lifespan.

## 1. Introduction

### 1.1. Preadolescent suicidality

Suicide is a complex behavior driven by complex environmental and genetic factors, and their interaction (Turecki and Brent, 2016). Suicide is the second leading cause of death in youth (Centers for Disease Control and Prevention, 2021), with steadily increasing rates of childhood and preadolescent suicidal ideation and suicide attempts (i.e., suicidality) particularly for Black-American youth (Bridge et al., 2018; Lindsey et al., 2019; Mishara and Stijelja, 2020; Naghavi and Global Burden of Disease Self-Harm, 2019; Plemmons et al., 2018; Ruch et al., 2019; Yu and Chen, 2019). Childhood suicidality predicts adult psychiatric morbidity and mortality and thus may represent an early marker for lifelong vulnerability to poor mental health (Copeland et al., 2017; Herba et al., 2007). Understanding risk factors of preadolescent suicidality is critical as it can guide prevention and intervention early in the lifespan (Cash and Bridge, 2009).

### 1.2. Genetics and suicidality

Genetic risk for suicidality is understudied, with the first genome wide association studies (GWAS) emerging recently (Mullins et al., 2019; Turecki et al., 2019), with no genetic studies of youth suicidality. One potential approach to better understand the genetic underpinning of suicidal behavior is to study polygenic risk for phenotypes associated with suicide, like neuropsychiatric risk (Bigdeli et al., 2020; Laursen et al., 2017; Warrier and Baron-Cohen, 2019). In light of the mounting evidence linking environmental adversity with suicidal behavior (Fazel and Runeson, 2020; Turecki et al., 2019), other predisposition factors (such as genetic risk) that also influence stress reactivity likely also contribute to suicidal behavior. Progress has been made in the understanding of the genetic underpinnings of stress susceptibility with the latest GWAS of post-traumatic-stress-disorder (PTSD) showing significant heritability (Gelernter et al., 2019; Nievergelt et al., 2019; Stein et al., 2021) with the involved genomic loci starting to reveal diverse biological pathways in brain and non-brain tissues and cell types (Chatzinakos et al., 2020; Girgenti et al., 2021; Huckins et al., 2020). GWAS also allow for calculation of a polygenic risk score (PRS) of a trait in a separate target population not used in the GWAS (Crouch and Bodmer, 2020). The PRS of a trait is expected to not only be predictive of that trait’s phenotype in the larger population, but also to be predictive of other related phenotypes (Shen et al., 2020). A major challenge in applying polygenic risk is to calculate a PRS that accounts for diversity among ancestries, with the vast majority of research excluding non-European ancestry individuals thus limiting their wider generalizability (Duncan et al., 2019; Martin et al., 2019).

### 1.3. ABCD Study® as a resource to study preadolescent suicidality

Here we aimed to address the gap in studies of genetic susceptibility to early life suicidal behavior, using data from the Adolescent Brain Cognitive Development study (ABCD Study®), which included a genotyped population of diverse US preadolescents with phenotyping of environmental adversities (Hoffman et al., 2019) and suicidality (Janiri et al., 2020). We calculated ancestry-specific PTSD-PRS for European ancestry and African ancestry participants from the ABCD Study®. We evaluated whether PTSD-PRS, as a genetic indicator of stress susceptibility, can predict preadolescent suicidality. We further evaluated whether PTSD-PRS has additive or interactive effects with early life adversities that are known risk factors associated with preadolescent suicidality. Our analytic approach was ancestry-specific, but we also meta-analyzed the two ancestries to estimate effects across ancestries. We hypothesized that (1) PTSD-PRS would be predictive of preadolescent suicidality, but with a smaller effect size than environmental adversity; and that (2) ancestry-specific PRS would outperform cross-ancestry PRS in the prediction of preadolescent suicidality.

## 2. Methods

### 2.1 Participants

The ABCD Study® sample includes a cohort of 11,878 children aged 9–10 years at baseline, primarily recruited through local school systems (Garavan et al., 2018). Participants were enrolled at 21 sites, with the catchment area encompassing over 20% of the entire US population in this age group. All participants gave informed assent. Parents/caregivers signed informed consent. The ABCD Study® protocol was approved by the University of California, San Diego Institutional Review Board (IRB), and was exempted from a full review by the University of Pennsylvania IRB.

Here we used data collected at the 1-year follow-up assessment, which included a tool that assessed self-reported experiences of adversity including negative life events and experiences of discrimination. In some cases, variables were only available from baseline assessment, either because measures were deemed non-longitudinal, or because the ABCD Study® data collection schedule did not include these items at the 1-year follow-up (see **supplementary Table S1** for details on all ABCD Study® measures used in the current analysis). We included phenotypic data from ABCD Study® data release 3.0 (https://abcdstudy.org/), while PRS were calculated using genotype data from release 2.0.1. To mitigate potential effects of family-relatedness, as ABCD Study® included twins and other related individuals, we only included one participant per family (the oldest). The final sample analyzed here included participants from European ancestry (EUR, N=4,619) and from African ancestry (AFR, N=1,334), based on inferred genetic ancestry rather than on parental reports of race (detailed below).

### 2.2. Calculation of ancestry-specific PTSD-PRS

PRS are commonly computed using summary statistics from GWAS of independent population(s) from the same ancestry group. Since the vast majority of GWAS have been run only for EUR samples, computing PRS for non-EUR samples often forces researchers to rely on EUR GWAS, with limited evidence to support this approach (Duncan et al., 2019; Martin et al., 2019). Using GWAS summary statistics from the Psychiatric Genomics Consortium-Posttraumatic Stress Disorder Group (PGC-PTSD) that were computed separately for EUR (N = 23,212 cases and 151,447 controls) and AFR (N = 4,363 cases and 10,976 controls) (Nievergelt et al., 2019), we calculated ancestry specific PTSD-PRS for EUR and AFR participants of the ABCD Study®, as described below.

Participants from the ABCD Study® provided biospecimens (Uban et al., 2018), and genotype data were obtained from NDA (#2573, fix release 2.0.1). Prior to imputation, PLINK 1.9 (Chang et al., 2015) was used to remove single nucleotide polymorphisms (SNPs) with > 5% missingness, samples with more than 10% missingness, and samples in which genotyped sex was different from reported sex. Genotypes were phased (Eagle v.2.4) and imputed to the 1000 Genomes Project Other/Mixed GRCh37/hg19 reference panel (Phase 3 v.5) using Minimac 4 via the Michigan Imputation Server (Das et al., 2016). Following imputation, only polymorphic sites with imputation quality *R*^2^ ≥ 0.7 and MAF ≥ 0.01 were retained.

Multi-dimensional scaling (MDS) of the imputed genotypes was conducted using KING v.2.2.4 (Manichaikul et al., 2010) to identify the top ten ancestry principal components (PCs) for each sample. These PCs were projected onto the 1000 Genomes Project PC space, and genetic ancestry was inferred using the e1071 (Meyer et al., 2020) support vector machines package in R (R Core Team, 2019) (**Supplementary Figure S1**). Based on these inferences, AFR and EUR cohorts were established for the ABCD Study® dataset; all other ancestry groups were excluded from further analysis. A second round of unprojected MDS was then performed within the EUR and AFR groups to produce ten PCs that were regressed out of the standardized PRS to adjust for genetic ancestry (**Supplementary Figures S2-S3**). KING was also used to identify all pairwise relationships out to third degree relatives based on estimated kinship coefficients and inferred IBD segments.

PRS-CS (PRS using SNP effect sizes under continuous shrinkage) (Ge et al., 2019) was used to infer posterior effect sizes of the SNPs in the dataset that overlapped with the PGC-PTSD GWAS summary statistics and an external 1000 Genomes linkage disequilibrium (LD) panel matched to the ancestry group used for the GWAS. PRS for the EUR and AFR subsets of the ABCD Study® participants were computed twice, once using the EUR LD panel and EUR GWAS and once using the AFR LD panel and AFR GWAS. Raw PRS were produced by PLINK 1.9 and then standardized and ancestry-corrected in R.

### 2.3. Phenotypic measures

#### 2.3.1. Exposures

The ABCD Study® assessment included multiple instruments that tapped adversity. For the current analysis, we specifically chose measures that had been previously linked to increased suicide risk in youth: negative life events (a continuous count of events judged as “bad” by the study’s participants, measure ple_y_ss_total_bad) (Serafini et al., 2015); family conflict (child-reported sub-scale from the Family Environment Scale, measure fes02fes_y_ss_fc) (Janiri et al., 2020); and discrimination against lesbian, gay, or bisexual, (LGB) individuals (past 12-months, binary variable, measure dim_yesno_q3) (Almeida et al., 2009).

#### 2.3.2. Outcome measures

The ABCD Study® included the Kiddie-Structured Assessment for Affective Disorders and Schizophrenia for DSM-5 (KSADS-5) (Kaufman et al., 1997) that assessed past or current suicidal ideation and attempt (Janiri et al., 2020). Items relating to self-injurious behavior without suicidal intent were not included. As the proportion of suicidal attempts was low, and to avoid multiple testing to mitigate risk of type I error, we grouped together suicidal ideation and attempt, in line with previous analyses (Janiri et al., 2020). Thus, suicidal outcomes were collapsed into a single binary measure termed *suicidality*.

### 2.4. Statistical Analyses

We used the SPSS 26.0 statistical package and R 3.6.1 for our data analysis. Mean (standard deviation [SD]) and frequency (%) were reported for descriptive purposes. Univariate comparisons were made using t-tests for continuous measures and chi-square tests for categorical variables. We employed listwise deletion for participants with missing data. Rates of missing values for all variables included in the current study were lower than 1.7%, with the exception of LGB discrimination that was missing in 5.1% in European Ancestry and 7.4% in the African Ancestry participants. We used two-tailed tests for all models. Data analyses were conducted between January and February 2021. Data preprocessing and analyses are detailed at https://github.com/barzilab1/abcd_discrimination.

#### 2.4.1. Main analysis

To determine the association of PTSD-PRS with preadolescent suicidality, we conducted a binary logistic regression model with the PTSD-PRS (residualized using 10 ancestry PCs) as the independent variable and suicidality (binary) as the dependent variable, co-varying for age and sex. To allow ancestry specificity, we used a PRS computed in a EUR GWAS training set for the EUR participants, and a PRS computed in an AFR GWAS training set for the AFR participants. We visualized the association between PTSD-PRS in both ancestries in both sexes across PRS quintiles (**Figure 1**). To estimate the effect of PRS in the AFR and EUR samples combined, we conducted fixed effect inverse-variance meta-analyses using the ancestry specific PRS, using the following formula: 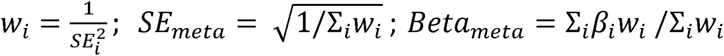, where *β*_*i*_ is the standardized regression coefficient of the PRS for the two ancestry groups (denoted by *i*), *SE*_*i*_ is the associated standard error, and *w*_*i*_ is the weight. P-values were calculated from Z scores (*Beta*_*meta*_/*SE*_*meta*_). We report Nagelkerke’s R^2^ for each ancestry separately but not for the meta-analysed results as there is currently no consensus about the best method to calculate this.

**Figure 1.**
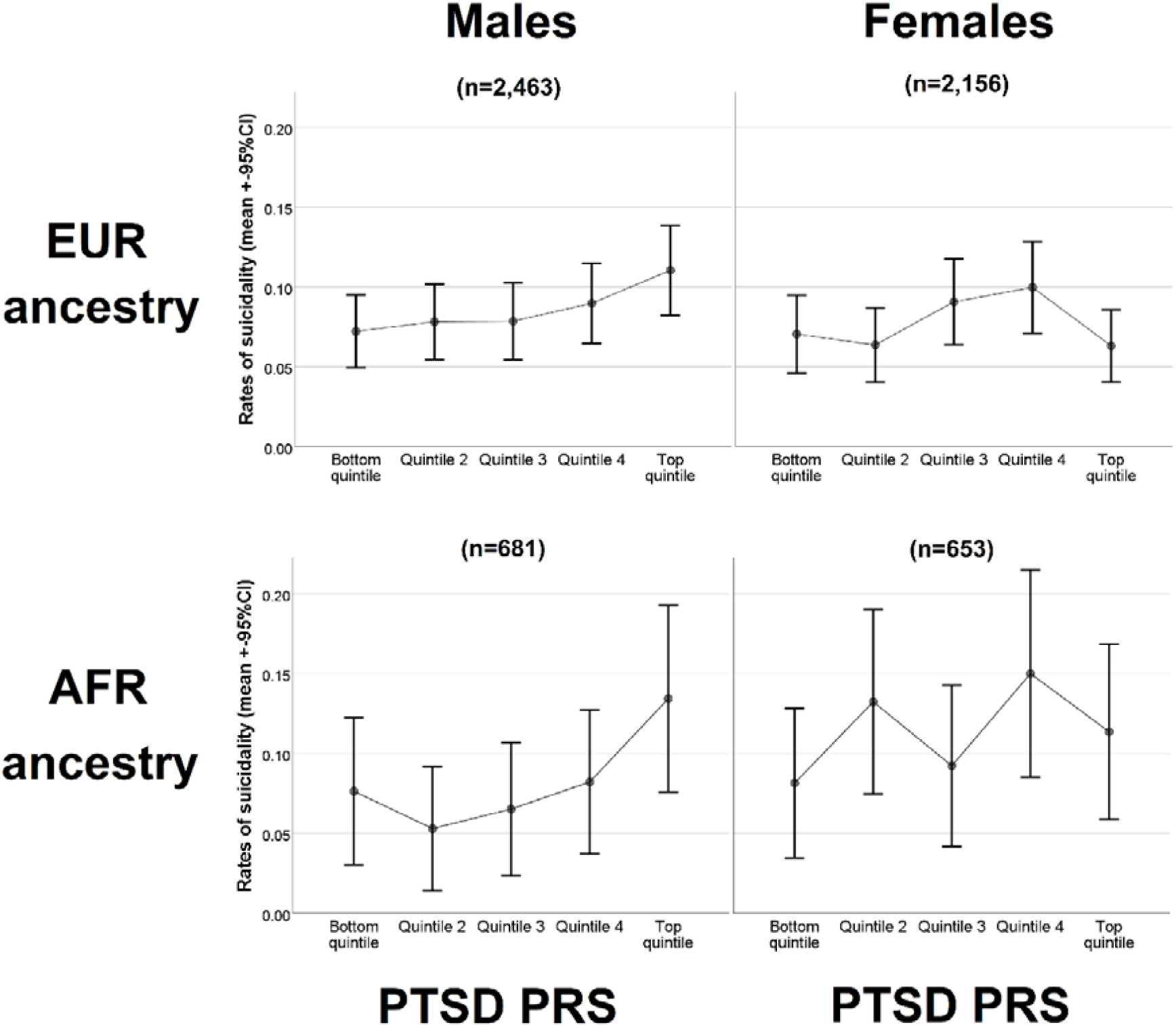
Visual representation of ancestry-specific PTSD-PRS and childhood suicidality by ancestry and sex. Abbreviations: EUR= European ancestry; AFR= African Ancestry; PTSD= post-traumatic stress disorder; PRS= polygenic risk score.

To estimate environmental adversities’ effects in participants from each ancestry (EUR and AFR), we conducted a binary logistic regression model with the three environmental adversities (negative life events, family conflict, and LGB discrimination) as the independent variables and suicidality (binary) as the dependent variable, co-varying for age and sex. Additionally, we conducted fixed effect inverse-variance weighted meta-analysis of the EUR and AFR samples combined for each of the three environmental adversities.

To estimate whether PTSD-PRS can additively explain variability in suicidality over and above environmental adversities, we used a stepwise approach. We first ran binary logistic regression models with each of the environmental adversities as the independent variables, and suicidality as the dependent variable (Step 1). Thereafter, we added the PTSD-PRS (Step 2) to evaluate whether the model explains more variability (i.e., increase in the R-square). Lastly, we tested whether there is evidence for G X E through the introduction of the environmental adversity X PTSD-PRS product (Step 3). This stepwise approach was done separately for each of the three environmental adversities (and not in a single model), to allow maximal chance of PTSD-PRS to explain variability. For steps 1 and 2, we conducted fixed effect inverse-variance weighted meta-analysis of the EUR and AFR samples combined for each of the three environmental adversities.

#### 2.4.2. Sensitivity Analyses

To test the specificity of the ancestry-specific PTSD-PRS to suicidality we ran two models where we substituted the ancestry-specific PTSD-PRS. In the first model, we included an ancestry-specific polygenic score for a non-related phenotype, height PRS (using a diverse training set that allowed calculation of EUR and AFR PRS (Marouli et al., 2017)). This model allowed us to test whether the PTSD-PRS is specific to the psychiatric phenotype. In the second model, we included the cross-ancestry PTSD-PRS, so that in EUR participants we used the AFR derived PTSD-PRS, and vice versa. This allowed us to test whether the ancestry specificity is important when testing the association between PTSD-PRS and suicidality and to assess whether PTSD-PRS calculated using the larger, more powerful EUR PTSD GWAS had the potential to explain more variance in the AFR sample than PTSD-PRS from a smaller, but ancestry-matched, GWAS.

## 3. Results

### 3.1. Comparison between participants with and without suicidality

Comparison of demographics, PTSD-PRS and experiences of environmental adversities between Suicidal and Non-suicidal participants is described in **Table 1**. Participants who endorsed suicidality comprised 8.2% of the EUR participants and 9.7% of the AFR participants. Univariate comparison did not reveal significant age or sex differences. As expected, participants who endorsed suicidality had more experiences of all three environmental adversities, in both ancestries (all p’s<0.001).

**Table 1.** Sample characteristics

|  |  | EUR sample (N=4,619) |  |  |  |  | AFR sample (N=1,334) |  |  |  |  |
| --- | --- | --- | --- | --- | --- | --- | --- | --- | --- | --- | --- |
|  |  | No suicidality<br>(n=4,241) |  | Suicidality<br>(n=378) |  | p-value | No suicidality<br>(n=1,204) |  | Suicidality<br>(n=130) |  | p-value |
|  |  | Mean/n | SD/% | Mean/n | SD/% |  | Mean/n | SD/% | Mean/n | SD/% |  |
| **Demographics** | Age years, mean (SD) | 10.99 | 0.63 | 11 | 0.65 | 0.61 | 10.95 | 0.63 | 11 | 0.58 | 0.362 |
|  | Sex male, n (%) | 2,252 | 53.10% | 211 | 55.80% | 0.31 | 625 | 51.90% | 56 | 43.10% | 0.056 |
|  | Sex female, n (%) | 1,989 | 46.90% | 167 | 44.20% | 0.31 | 579 | 48.10% | 74 | 56.90% | 0.056 |
| **Ancestry-specific** | PTSD-PRS, mean (SD) | -0.01 | 0.96 | 0.10 | 0.93 | 0.04 | -0.01 | 0.7 | 0.08 | 0.64 | 0.16 |
| **Genomic scores** | Height PRS, mean (SD) | 0 | 0.95 | -0.07 | 0.94 | 0.14 | 0.01 | 0.95 | -0.01 | 0.96 | 0.88 |
| **Environmental**<br>**adversities** | Negative life events count, mean (SD) | 2.09 | 1.97 | 3.48 | 2.69 | <.001 | 3.05 | 2.61 | 4.92 | 2.92 | <.001 |
|  | Family conflict scale, mean (SD) | 1.64 | 1.77 | 2.8 | 2.12 | <.001 | 2.12 | 1.83 | 2.85 | 2.1 | <.001 |
|  | LGB discrimination, n (%) | 107 | 2.52% | 52 | 13.76% | <.001 | 53 | 4.40% | 22 | 16.92% | <.001 |

Abbreviations: EUR= European ancestry; AFR= African ancestry; SD= standard deviation; PTSD= post-traumatic stress disorder; LGB= Lesbian, gay, bisexual.

### 3.2. Association of ancestry-specific PTSD-PRS and suicidality

In EUR participants, a PTSD-PRS calculated from a EUR GWAS (i.e., ancestry-specific) was significantly associated with suicidality with a modest effect size (odds ratio [OR]=1.12, 95%CI=1.01-1.25, p=0.038), with Nagelkerke R^2^ =0.003. In AFR participants, the AFR PTSD-PRS was not significantly associated with suicidality, but a similar strength and direction of effect was observed (OR=1.21, 95%CI 0.93-1.57, p=0.153), with Nagelkerke R^2^ =0.010. No differences in the height polygenic scores were observed between suicidal and non-suicidal participants in both ancestries.

Furthermore, meta-analysis of the AFR and EUR samples identified significant effects (OR = 1.14, 95%CI 1.03-1.26, p=0.014), possibly indicating that AFR analysis was underpowered. **Table 2** details full model statistics for both ancestries. **Figure 1** presents a visual presentation of the relationship between PTSD-PRS and suicidality in both ancestries and both sexes.

**Table 2.**
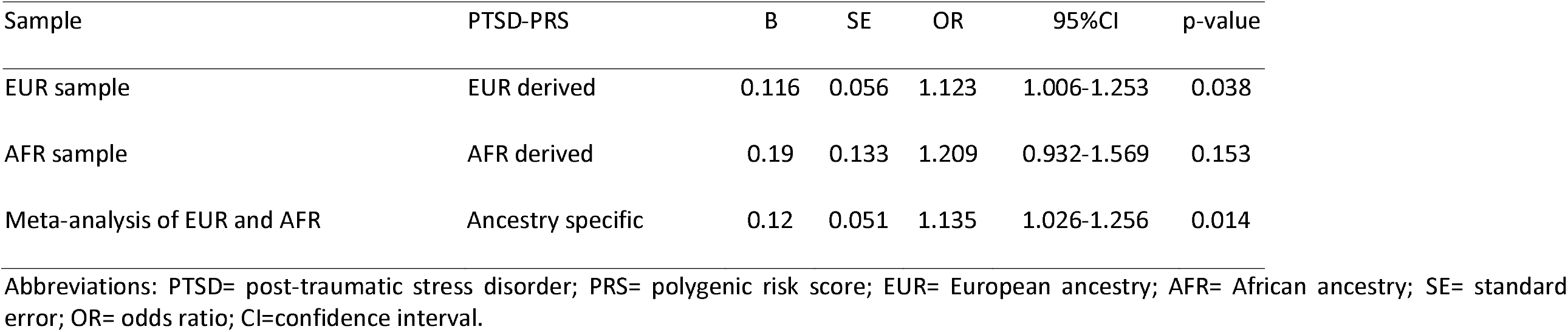
Ancestry specific PTSD-PRS’ association with childhood suicidality

We then conducted a sensitivity analysis to assess specificity of the finding of a significant association of the ancestry-specific PTSD-PRS with suicidality in the EUR participants (**Supplementary Table S2**). Substitution of the PTSD-PRS with a PRS for non-psychiatric phenotype (polygenic score for height) was not associated with suicidality; nor was a cross-ancestry PTSD-PRS derived from an AFR population, supporting specificity for the ancestry-specific PTSD-PRS effect on suicidality.

### 3.3. Environmental adversity and suicidality

In both ancestries, environmental adversities were significantly associated with suicidality with small-to-moderate effect size for negative life events and family conflict (OR ranging from 1.27-1.6 in EUR, AFR, and EUR/AFR meta-analytic samples); and with large effect size (OR ∼4.0 in EUR, AFR, and EUR/AFR meta-analytic samples) for LGB discrimination (**Table 3**).

**Table 3.**
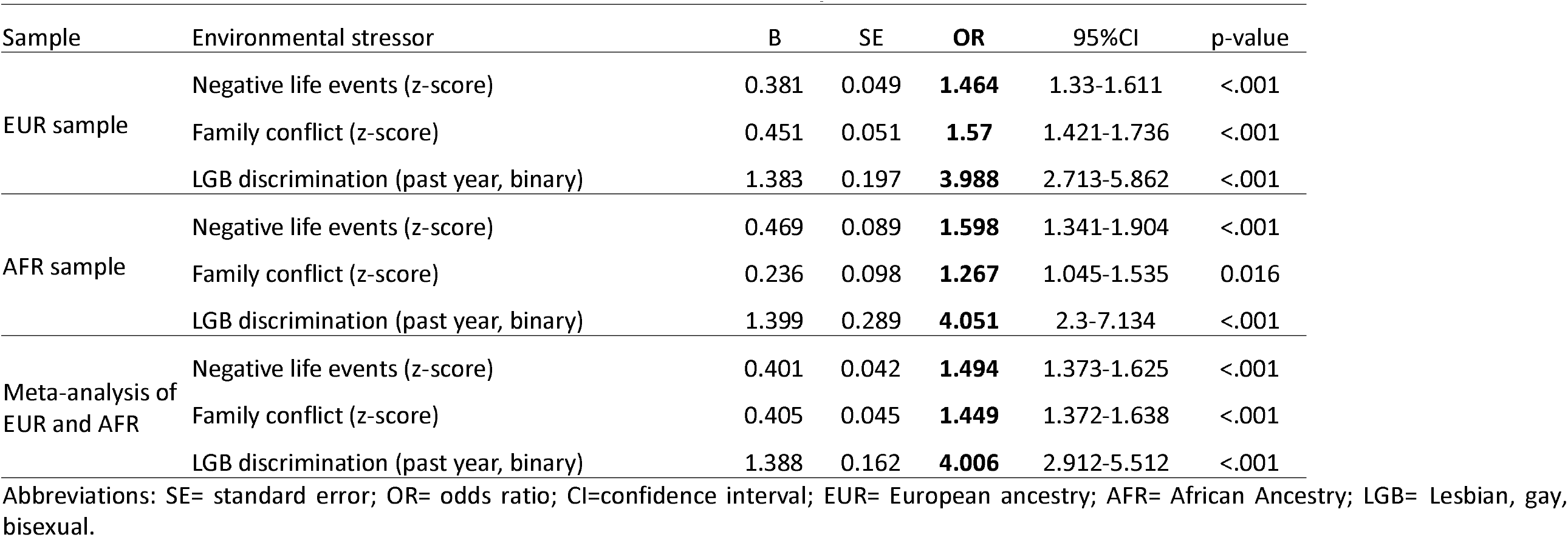
Associations of environmental adversities with childhood suicidality

| Sample | Environmental stressor | B | SE | OR | 95%CI | p-value |
| --- | --- | --- | --- | --- | --- | --- |
| EUR sample | Negative life events (z-score) | 0.381 | 0.049 | **1.464** | 1.33-1.611 | <.001 |
|  | Family conflict (z-score) | 0.451 | 0.051 | **1.57** | 1.421-1.736 | <.001 |
|  | LGB discrimination (past year, binary) | 1.383 | 0.197 | **3.988** | 2.713-5.862 | <.001 |
| AFR sample | Negative life events (z-score) | 0.469 | 0.089 | **1.598** | 1.341-1.904 | <.001 |
|  | Family conflict (z-score) | 0.236 | 0.098 | **1.267** | 1.045-1.535 | 0.016 |
|  | LGB discrimination (past year, binary) | 1.399 | 0.289 | **4.051** | 2.3-7.134 | <.001 |
| Meta-analysis of<br>EUR and AFR | Negative life events (z-score) | 0.401 | 0.042 | **1.494** | 1.373-1.625 | <.001 |
|  | Family conflict (z-score) | 0.405 | 0.045 | **1.449** | 1.372-1.638 | <.001 |
|  | LGB discrimination (past year, binary) | 1.388 | 0.162 | **4.006** | 2.912-5.512 | <.001 |

Abbreviations: SE= standard error; OR= odds ratio; CI=confidence interval; EUR= European ancestry; AFR= African Ancestry; LGB= Lesbian, gay, bisexual.

### 3.4. Integrating PTSD polygenic risk into environmental models of suicidality

Lastly, we tested the contribution of PTSD-PRS to explaining variability in suicidality over and above environmental adversity. The addition of PTSD-PRS made a small additive contribution to each model including the one of the adversity measures (negative life events, family conflict, or LGB discrimination) (**Table 4**). While the addition of PTSD-PRS effects was not significant in any of the three adversity models in each of either EUR or AFR ancestry-specific analyses separately, meta-analyses of the EUR and AFR samples identified a significant effect of PTSD-PRS after accounting for negative life events and family conflict, but not LGB discrimination (**Table 4**). No evidence for G X E were observed (for each adversity X PTSD-PRS, p’s>0.38).

**Table 4.** Additive effects of PTSD-PRS in explaining childhood suicidality using environmental adversities

| EUR participants (N=4,619) |  |  |  |  |  |  |  |  |  |
| --- | --- | --- | --- | --- | --- | --- | --- | --- | --- |
|  | Model 1 (E effect) |  |  | Model 2 (E effect + PTSD-PRS (G)) |  |  | Model 3 (E, G + G X E) |  |  |
|  | OR | 95%CI | p | OR | 95%CI | p | OR | 95%CI | p |
| Negative life events (z) | 1.64 | 1.51-1.79 | <.001 | 1.64 | 1.501-1.78 | <.001 | 1.64 | 1.51-1.79 | <.001 |
| PTSD-PRS |  |  |  | 1.1 | 0.99-1.23 | 0.091 | 1.14 | 0.96-1.35 | 0.139 |
| G X E |  |  |  |  |  |  | 0.99 | 0.95-1.03 | 0.609 |
| Nagelkerke R<sup>2</sup> | 0.061 |  |  | 0.063 |  |  | 0.063 |  |  |
| Family conflict (z) | 1.67 | 1.53-1.82 | <.001 | 1.67 | 1.53-1.82 | <.001 | 1.66 | 1.52-1.82 | <.001 |
| PTSD-PRS |  |  |  | 1.12 | 1-1.25 | 0.055 | 1.07 | 0.9-1.27 | 0.462 |
| G X E |  |  |  |  |  |  | 1.02 | 0.97-1.07 | 0.531 |
| Nagelkerke R<sup>2</sup> | 0.059 |  |  | 0.061 |  |  | 0.061 |  |  |
| LGB discrimination (binary) | 6.97 | 4.71-9.53 | <.001 | 6.66 | 4.68-9.48 | <.001 | 6.74 | 4.74-9.6 | <.001 |
| PTSD-PRS |  |  |  | 1.09 | 0.97-1.22 | 0.156 | 1.11 | 0.98-1.26 | 0.102 |
| G X E |  |  |  |  |  |  | 0.85 | 0.59-1.22 | 0.383 |
| Nagelkerke R<sup>2</sup> | 0.048 |  |  | 0.049 |  |  | 0.049 |  |  |
| AFR participants (N=1,334) |  |  |  |  |  |  |  |  |  |
|  | Model 1 (E effect) |  |  | Model 2 (E effect + PTSD-PRS (G)) |  |  | Model 3 (E, G + G X E) |  |  |
|  | OR | 95%CI | p | OR | 95%CI | p | OR | 95%CI | p |
| Negative life events (z) | 1.67 | 1.42-1.96 | <.001 | 1.67 | 1.42-1.96 | <.001 | 1.66 | 1.42-1.96 | <.001 |
| PTSD-PRS |  |  |  | 1.2 | 0.92-1.57 | 0.178 | 0.87 | 0.54-1.39 | 0.558 |
| G X E |  |  |  |  |  |  | 1.08 | 0.99-1.18 | 0.101 |
| Nagelkerke R<sup>2</sup> | 0.065 |  |  | 0.068 |  |  | 0.072 |  |  |
| Family conflict (z) | 1.45 | 1.23-1.71 | <.001 | 1.45 | 1.23-1.71 | <.001 | 1.45 | 1.22-1.71 | <.001 |
| PTSD-PRS |  |  |  | 1.21 | 0.93-1.67 | 0.158 | 1.19 | 0.77-1.83 | 0.433 |
| G X E |  |  |  |  |  |  | 1.01 | 0.89-1.14 | 0.927 |
| Nagelkerke R<sup>2</sup> | 0.036 |  |  | 0.039 |  |  | 0.039 |  |  |
| LGB discrimination (binary) | 4.84 | 2.8-8.34 | <.001 | 4.87 | 2.82-8.41 | <.001 | 4.83 | 2.79-8.37 | <.001 |
| PTSD-PRS |  |  |  | 1.23 | 0.93-1.64 | 0.145 | 1.2 | 0.89-1.64 | 0.236 |
| G X E |  |  |  |  |  |  | 1.18 | 0.53-2.64 | 0.68 |
| Nagelkerke R<sup>2</sup> | 0.054 |  |  | 0.058 |  |  | 0.058 |  |  |
| Meta-analysis of EUR and AFR (N=5,953) |  |  |  |  |  |  |  |  |  |
|  | Model 1 (E effect) |  |  | Model 2 (E effect + PTSD-PRS (G)) |  |  |  |  |  |
|  | OR | 95%CI | p | OR | 95%CI | p |  |  |  |
| Negative life events (z) | 1.647 | 1.53 - 1.78 | <.001 | 1.642 | 1.524 - 1.769 | <.001 |  |  |  |
| PTSD-PRS |  |  |  | 1.115 | 1.006 - 1.236 | 0.038 |  |  |  |
| Family conflict (z) | 1.616 | 1.52 - 1.78 | <.001 | 1.614 | 1.491 - 1.747 | <.001 |  |  |  |
| PTSD-PRS |  |  |  | 1.128 | 1.073 - 1.186 | 0.021 |  |  |  |
| LGB discrimination (binary) | 6.084 | 4.53 - 8.18 | <.001 | 6.073 | 4.516 - 8.169 | <.001 |  |  |  |
| PTSD-PRS |  |  |  | 1.109 | 1.051 - 1.169 | 0.061 |  |  |  |

## 4. Discussion

### 4.1. Broad context

In this study, we investigated the effects of stress-related genetic factors and stress exposures in relation to the development of preadolescent suicidality. We linked PRS of stress susceptibility (i.e., PRS of PTSD) with preadolescent suicidality, indicating a genetic overlap in this young population that has so far received limited attention in the context of genetic risk for suicidal ideation or attempts. Our findings add to the existing literature in adult populations linking other psychiatric PRS (for depression, schizophrenia and autism) to suicidal behavior (Bigdeli et al., 2020; Laursen et al., 2017; Warrier and Baron-Cohen, 2019). Although causal analysis need to be conducted, the observed influence of both environmental stressors and genetic stress-susceptibility on suicidality so early in the lifespan suggests that early life stress biology mechanisms are putative drivers of adulthood suicidal behavior and might be potential targets of suicide prevention approaches.

Our findings are important for several reasons. First, we show for the first time that a genetic marker of stress susceptibility derived from an adult psychiatric trait GWAS is associated with a clinical psychiatric phenotype in preadolescents. Notably, rate of preadolescent suicidality is showing an alarming rise (Ruch et al., 2019), and its genetic correlates are understudied. Second, we show that a PRS derived from a psychiatric disorder that is not suicide is relevant in explaining variability in suicidality even after accounting for environmental stressors, supporting the notion of using PRS across clinical phenotypes. In light of limited evidence on PRS of suicidal behavior, the current study supports further investigation into a cross-phenotype approach. Third, a key part of this analysis is the inclusion of AFR individuals in our PRS analyses. Although adversities are common across diverse populations and pose risk to the development for developing psychopathology, Black individuals face disproportionate exposure to adverse life events that contribute to such psychopathology (Slopen et al., 2016). Yet, psychiatric genetic research is skewed toward European ancestry groups (Duncan et al., 2019; Martin et al., 2019) which limits the generalizability of translational research towards the larger population. Our ancestry-specific design is especially important in light of recent epidemiological data showing that suicide rates in African American youth are disproportionately rising (Bridge et al., 2018; Price and Khubchandani, 2019), highlighting the critical need to study suicide in this population. Further inclusion of racial minorities in psychiatric genomic studies in the context of G X E is therefore critical for accurate understanding of stress biological mechanisms that lead to mental health dysfunction and the identification of robust PRS for prediction of psychopathology.

Suicide risk is modeled in a conceptual framework building on distal/predisposing risk, chronic environmental risk factors like lack of social cohesion and access to means, and proximal/precipitating factors, all leading to suicidal outcome (Turecki and Brent, 2016). This framework can be linked to the 3-hit hypothesis of stress-related developmental psychopathology, where a combination of genetic risk (hit-1) and early life stress (hit-2) can contribute to the expression of maladaptive behaviors after additional stress later in the lifespan (hit-3). This 3-hit hypothesis was originally coined as an interpretation of 55 years (1957-2012) of rodent and non-human primate literature of developmental programming by stress, and was supported by recent large scale cohorts (Daskalakis et al., 2013). In the current study, we leveraged the deep phenotyping of the ABCD Study and the genotyped sample to (1) model preadolescent suicidality along this framework, and (2) evaluate the effects of PTSD polygenic risk (hit-1) and early life environmental insults (hit-2) on suicidal behavior in preadolescents. Notably, we were able to show a significant additive effect of the PTSD-PRS genetic effects on suicidality risk even after accounting for environmental stress, though the conditional effects were attenuated compared to the marginal effects. We suggest that future work using longitudinal data (soon to be released), which will include real-time data collection through wearable and mobile technologies, can clarify the role of proximal risk factors and triggers for suicidal behavior (hit-3). Additionally, future longitudinal analyses of ABCD data that will include more granular life events (e.g., data on bullying that has not yet been released) might reveal G X E or E X E effects that were not identified in the current analysis.

### 4.2. Implications for suicide research

Our findings have two main implications for suicide research, (1) supporting the use of cross-phenotype genetic risk to study childhood suicidality, and (2) supporting inclusion of non-European ancestries in assessment of genetic risk. Regarding clinical implications, while we are cautious in data interpretation, our findings can provide preliminary data informing potential future use of PRS for suicide risk stratification. Notably, the PRS’ effects on suicidality are relatively small compared to the large effect size of environmental stressors and were not statistically significant in the ancestry-specific analyses accounting for environmental stressors. However, PRS additive effects were statistically significant in the meta-analytic approach even when accounting for the environmental stressors, implying that the lack of statistical significance in each ancestry is likely due to underpowered sample size of both testing and training samples. Thus, our study provides a proof-of-concept supporting the approach of integrating genomic tools in suicide research modeling along with environmental exposures. Lastly, while the implications detailed above seem practical for next steps in suicide research, the very small effect size of the PRS provides further evidence that use of PRS alone is likely of low/no clinical utility. However, our findings suggesting additive effect of PRS on strong environmental suicide risk factors indicate that combining PRS with other risk factors might be relevant for future risk stratification, as was recently suggested (Murray et al., 2021). More research is needed on how to leverage the powerful computational methodology behind PRS calculation and PRS from additional relevant phenotypes to inform clinical care and clinical decision making (Fullerton and Nurnberger, 2019).

### 4.3. Limitations

Limitations of this study include the use of AFR PRS based on AFR GWAS of PTSD, which was smaller and less well powered compared to the EUR GWAS of PTSD (Nievergelt et al., 2019). Future work is needed to expand AFR GWAS of PTSD for more well-powered AFR PRS estimates. In addition, we chose as an outcome of this study a phenotype that combined suicidal ideation and suicide attempt, which is consistent with prior work (Janiri et al., 2020), but is likely less accurate compared to defining separate suicidal phenotypes that probably have distinct biological underpinnings. This delineation to ideation versus attempt will require larger samples that are likely to be older. Lastly, we analyzed data collected at a single time point. As the ABCD study dataset grows, longitudinal analyses will be permitted and allow future work to investigate the predictive value of PTSD-PRS on studies evaluating genetic contribution to trajectories of suicidality over time.

## 5. Conclusions

In conclusion, our results support use of cross-phenotype PRS, specifically stress-susceptibility, as a genetic marker for risk of suicidality early in life. These data further demonstrate that stress-susceptibility PRS are markers for suicidality across ancestry groups and may thus be robust for prediction of suicidality. Future studies leveraging the longitudinal data expected to be released from the ABCD Study could evaluate causal relationships and the generalizability of the effects to older subjects, as well as study sex differences that are likely to emerge as the study population enters into mid- and late-adolescence.

## Data Availability

Data used in the preparation of this article were obtained from the Adolescent Brain Cognitive DevelopmentSM (ABCD) Study (https://abcdstudy.org), held in the NIMH Data Archive (NDA).

## 8. Acknowledgement

Data used in the preparation of this article were obtained from the Adolescent Brain Cognitive Development^SM^ (ABCD) Study (https://abcdstudy.org), held in the NIMH Data Archive (NDA). This is a multisite, longitudinal study designed to recruit more than 10,000 children age 9-10 and follow them over 10 years into early adulthood. The ABCD Study® is supported by the National Institutes of Health and additional federal partners under award numbers U01DA041048, U01DA050989, U01DA051016, U01DA041022, U01DA051018, U01DA051037, U01DA050987, U01DA041174, U01DA041106, U01DA041117, U01DA041028, U01DA041134, U01DA050988, U01DA051039, U01DA041156, U01DA041025, U01DA041120, U01DA051038, U01DA041148, U01DA041093, U01DA041089, U24DA041123, U24DA041147. A full list of supporters is available at https://abcdstudy.org/federal-partners.html. A listing of participating sites and a complete listing of the study investigators can be found at https://abcdstudy.org/consortium_members/. ABCD consortium investigators designed and implemented the study and/or provided data but did not necessarily participate in analysis or writing of this report. This manuscript reflects the views of the authors and may not reflect the opinions or views of the NIH or ABCD consortium investigators.

